# Evidence of varicella zoster virus (VZV) reactivation in children with arterial ischemic stroke: Results of the VIPS II Study

**DOI:** 10.1101/2024.05.26.24307958

**Authors:** Heather J. Fullerton, Nancy K. Hills, Max Wintermark, Nomazulu Dlamini, Catherine Amlie-Lefond, Michael M. Dowling, Lori C. Jordan, Timothy J. Bernard, Neil R. Friedman, Mitchell S.V. Elkind, Charles Grose, the VIPS II Investigators

## Abstract

**Background:** Varicella zoster virus (VZV) has been associated with focal cerebral arteriopathy (FCA) and arterial ischemic stroke (AIS) in childhood. The Vascular effects of Infection in Pediatric Stroke (VIPS) II study aimed to examine this relationship in the modern era when most children in North America and Australia receive VZV vaccination with live, attenuated virus.

**Methods:** This 22-center prospective cohort study enrolled 205 children (28 days-18 years) with AIS (2017-2022), collected baseline [hyperacute (≤72 hours; n=194) and acute (4-6 days; n=181)] and convalescent (1-6 weeks; n=74) serum samples. Sites enrolled 95 stroke-free controls with single serum samples. A virology research laboratory measured VZV IgM and IgG titers by an in-house enzyme-linked immunosorbent assay (ELISA). Baseline IgG seropositivity indicated prior exposure (vaccination/infection) and elevated IgM titers indicated recent reactivation.

**Results:** Median (IQR) age was 11.6 (5.5-15.6) years for cases and 11.8 (6.8-15.3) years for controls. Baseline serologies indicated prior VZV exposure in 198 cases (97%) and all controls. Parents of cases reported VZV vaccination in 160 (78%) and remote chicken pox in three (1.4%). Twenty cases (9.8%) and three controls (3.1%) had serologic evidence of recent VZV reactivation (p=0.06); all had remote VZV exposure (vaccination in 19 cases and all controls) and all were asymptomatic. Recent VZV reactivation was seen in similar proportions in arteriopathic, cardioembolic, and idiopathic stroke. Of 32 cases of FCA, 4 (12.5%) had recent VZV reactivation, versus no cases of arterial dissection (n=10) or moyamoya (n=16).

**Conclusions:** Serologic evidence of recent VZV reactivation (≈1-6 weeks prior to stroke) was present in one in 10 cases of childhood AIS, including those without arteriopathy. Clinically silent VZV reactivation may be a childhood stroke trigger despite widespread vaccination. These cases could represent waning immunity with reactivation of either vaccine virus or wild-type virus after an unrecognized secondary VZV infection.

## Introduction

Childhood arterial ischemic stroke (AIS) occurs annually in at least at least 2.4 per 100,000 US children, and the majority of affected children are previously healthy.^1, 2^ The pathogenesis of childhood AIS appears multifactorial, representing some combination of chronic and acquired risk factors. The “<u>V</u>ascular effects of Infection in <u>P</u>ediatric <u>S</u>troke” (VIPS I) Study demonstrated that common, minor childhood infections—mostly upper respiratory infections—transiently increase stroke risk for a matter of days.^3^ The mechanisms promoting AIS likely relate to circulating inflammatory cytokines affecting the coagulation system and endothelium (cardiac or arterial), as well as direct viral effects on endothelium.^4^

While this association between infection and AIS is not specific to any one virus, varicella zoster virus (VZV) may play a unique role in stroke pathogenesis. In the VIPS I study (2010-2014), 11.3% of 326 childhood AIS cases (versus 3% of 115 trauma controls), had positive VZV IgM antibodies on serum samples collected up to 3 weeks post-stroke, while VZV vaccination protected against stroke.^5^ VZV has been particularly linked to childhood focal cerebral arteriopathy (FCA), an acute, inflammatory stenotic disease typically involving a unilateral internal carotid artery (ICA) and its proximal branches.^6, 7^ FCA after chicken pox is also known as “post-varicella arteriopathy.”^8^ Because VZV remains latent in the trigeminal ganglion, and the trigeminal nerve innervates the cerebral arteries affected in FCA, VZV latent in trigeminal nerve is hypothesized to reactivate, travel antegrade along nerve branches to cerebral arteries, and result in a unilateral focal inflammatory arteriopathy.^9, 10^

However, the VZV landscape has changed in recent decades with most children in North America and Australia now receiving VZV vaccination in early childhood and chicken pox thus occurring infrequently. In addition, an alternate hypothesis exists for the relationship between VZV and stroke: rather than VZV triggering stroke, stroke may trigger VZV reactivation during the period of post-stroke down-regulation of immunity.^11^ The original VIPS study could not exclude this possibility due to the broad window for serum sample collection. In the VIPS II study, we aimed to examine the relationship between VZV and childhood AIS in the post-vaccination era and to address this question of causality in a new prospectively enrolled cohort with serum sample collection in earlier and narrower windows. We hypothesized that VZV infection—whether primary infection or reactivation of latent virus—precedes and triggers childhood AIS, and that VZV infection more commonly precedes arteriopathic stroke, particularly FCA, compared to cardioembolic or idiopathic stroke.

## Methods

### Setting and Case Enrollment

VIPS II is a prospective cohort study that enrolled children with spontaneous arteriopathic, cardioembolic, or idiopathic AIS at 22 centers (21 North American, one Australian) over a 5-year enrollment period: 12/2016-1/2022. Each site obtained local IRB approval and informed consent/assent for each patient enrolled. Inclusion criteria were age 28 days through 18 years at stroke ictus; AIS defined as acute onset neurological deficits with corresponding acute infarcts in an arterial territory on brain imaging; enrolled within 72 hours of stroke ictus (or last seen normal); and stroke type classified as arteriopathic, cardioembolic, or idiopathic at the time of enrollment. VIPS II excluded strokes that were iatrogenic (e.g., post-surgical or chemotherapy-related) because infection was less likely to play a role in stroke pathogenesis. To reduce heterogeneity, it excluded strokes that did not fall into one of the primary three categories (arteriopathic, cardioembolic, or idiopathic) and would have been classified as “other” stroke subtype in VIPS I (28 of 355 VIPS I cases).^12^ Sites collected blood samples at four time points relative to the stroke ictus: hyperacute (≤72 hours), acute (4-6 days), convalescent (1-6 weeks), and chronic (2-12 months). They interviewed parents or guardians within one week of enrollment using the VIPS I standardized script modified to remove time-consuming medication exposure questions; it included questions about prior chicken pox and VZV vaccination.^13^ Sites performed detailed abstraction of medical records into case report forms in a central REDCap^®^ database.

### Etiologic Classification

Clinically obtained brain and vascular imaging was collected for central review. A pediatric stroke neurologist (HJF) and neuroradiologist (MW) independently reviewed all available baseline and follow-up imaging and clinical data to confirm the AIS and centrally classify the stroke type using the methods of the original VIPS study;^12, 14^ disagreements were resolved through discussion. The primary classification categorized cases into four mutually exclusive groups: definite arteriopathy, possible arteriopathy, cardioembolic, and idiopathic. The idiopathic category included cases with no or modest stroke risk factors, such as genetic thrombophilias or isolated patent foramen ovale. Definite arteriopathy cases were further categorized into arteriopathy subtypes; intracranial dissections involving the anterior circulation were designated as FCA-d (dissection subtype).^12^ The brain imaging was also used to measure the volume of infarcts (using ABC/2).^14^

### Control Enrollment

VIPS II also had a case-control study compenent, enrolling unmatched stroke-free outpatient well controls (aged 28 days-18 years) at 17 sites (as allowed during pandemic-era restrictions). “Well” was defined as no infection, acute hospitalization, or immunizations in the prior two weeks. After obtaining informed consent, sites collected blood samples at a single time point and performed the same standardized parental interview used in the cases.

### VZV Serologies

A varicella research laboratory used an established in-house ELISA (enzyme linked immunosorbent assay) to measure VZV IgM and IgG serologies on serum samples from each available time point.^15^ The antigen for the ELISA was the VZV glycoprotein, gE (previously called gpI); gE is the most abundant VZV glycoprotein and has no cross-reactivity with any herpes simplex virus (HSV) protein.^15, 16^ Anti-human secondary antibodies conjugated to alkaline phosphatase were purchased from Thermo Scientific (goat anti-human IgG, catalog # 62-8422, and goat anti-human IgM, # A18844). The plate was read at 405nm wavelength. A negative titer (no antibody detected) was considered a level ≤0.1 absorbance units. The VZV antibody assays do not distinguish between wild-type VZV and the live, attenuated VZV in the chicken pox vaccine. All assays were carried out in the same laboratory with the same equipment under the direction of the same laboratory supervisor.

### Definitions of VZV Infection

For “baseline serologies,” we used the hyperacute and acute blood samples, while “convalescent serologies” were based on the convalescent sample, or chronic sample if no convalescent sample was available. A pediatric virologist (CG) interpreted the results and classified the evidence of infection. In general, with a primary VZV infection (chicken pox), IgM and IgG antibodies appear within 2-5 days of the rash.^17–19^ IgM antibodies peak at 2-3 weeks, then rapidly drop, usually becoming undetectable within 3-6 weeks; IgG antibodies wane but remain present. With reactivation (zoster), IgM antibodies are again produced, followed by a rise in IgG antibodies (“anamnestic response”). In studies of shingles (herpes zoster), IgM levels are variably detectable within 3-7 days of the rash, peak at 7-30 days (at lower levels than after chicken pox), and become undetectable within 4-12 weeks. ^17–19^ We defined serologic evidence of <u>prior exposure</u> (infection or vaccination) as baseline IgG titers >0.1 absorbance units; low-level IgG titers (>0.1 and <1.1) were consistent with prior exposure and no recent infection (reactivation). Serological evidence of <u>recent infection (≈ 1-6 weeks prior)</u> at the time of stroke (or control enrollment) was defined as a baseline IgM titer ≥1.1 (highest titer used if two baseline samples). Recent infection with baseline IgG≤0.1 would indicate primary infection, while recent infection with baseline IgG >0.1 indicated <u>reactivation</u>. Baseline IgG titers >1.1 provide additional evidence of recent reactivation (anamnestic response). Serological evidence of *<u>post</u>*<u>-stroke reactivation</u> was defined as a baseline IgG >0.1 and <1.1, baseline IgM titer <1.1, and either a 4-fold rise in IgG titers (baseline to convalescent) or a convalescent IgM titer ≥1.1.

### Data Analysis

Baseline characteristics of cases with versus without recent VZV reactivation were compared using Wilcoxon rank sum tests for continuous variables and chi-square tests (or Fisher’s exact, when appropriate) for categorical variables. All analyses were conducted using Stata v17.0 (Stata Corp., College Station, TX) with alpha set at 0.05. The p-values are two-sided. We completed the STROBE checklist for observational studies.

## Results

The 22 sites enrolled 205 cases of spontaneous AIS: 188 in the US, 15 in Canada, and 2 in Australia. Cases were 45% female and racially diverse with a median (IQR) age at stroke ictus of 11.6 (5.5-15.6) years (**Table 1**). Upon central classification, the stroke subtypes included 75 (37%) definite arteriopathy, 78 (38%) possible arteriopathy, 24 (12%) cardioembolic, and 38 (14%) idiopathic. Seventeen sites enrolled 100 well controls (95 with blood samples available for VZV serologies). Median (IQR) age of the 95 controls with blood samples was 11.8 (6.8-15.3) years. Control age and sex were similar to cases, but controls were more likely to be of mixed or other race and had higher socioeconomic status (**Supplemental Table 1**).

**Table 1.**
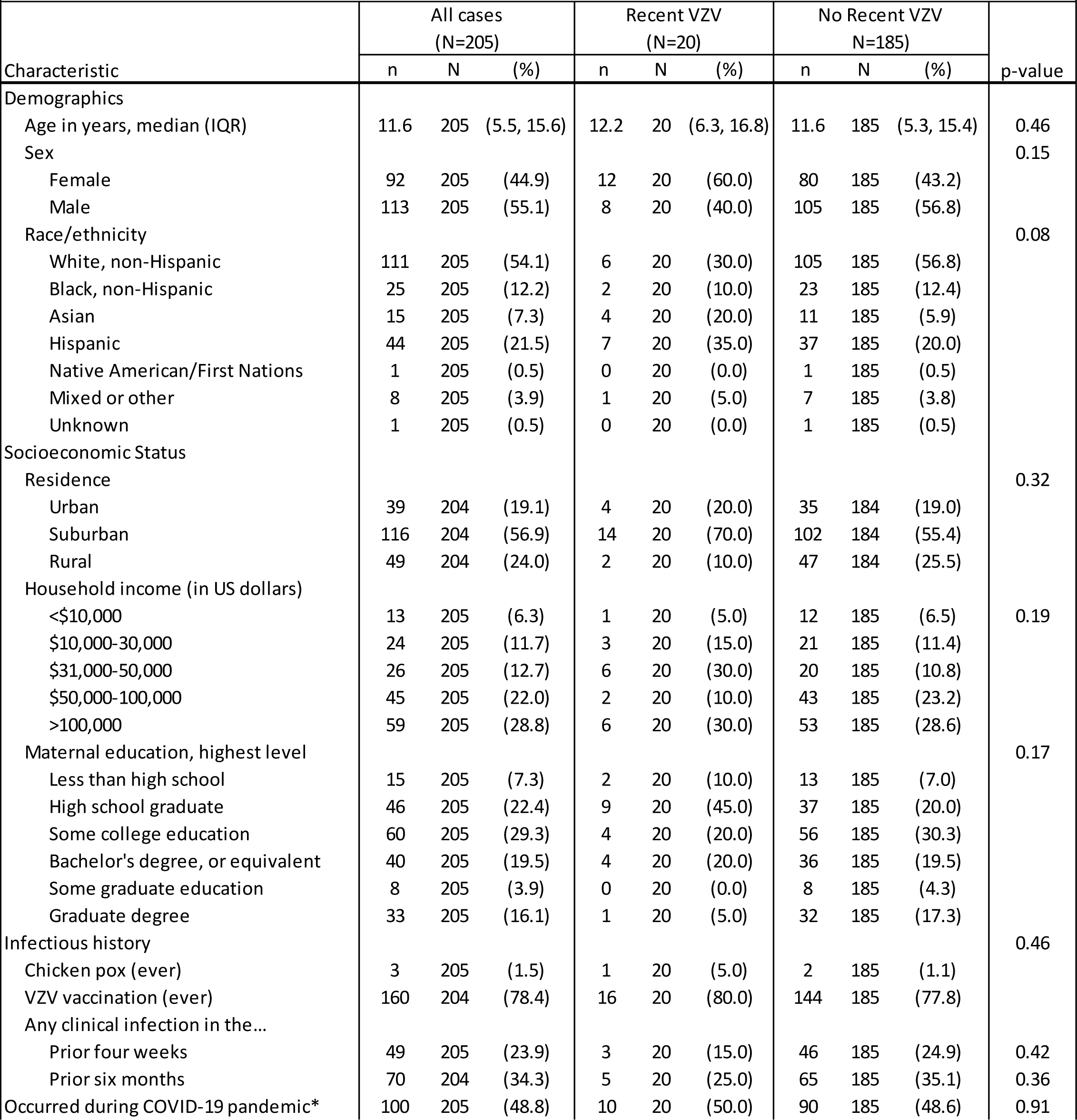
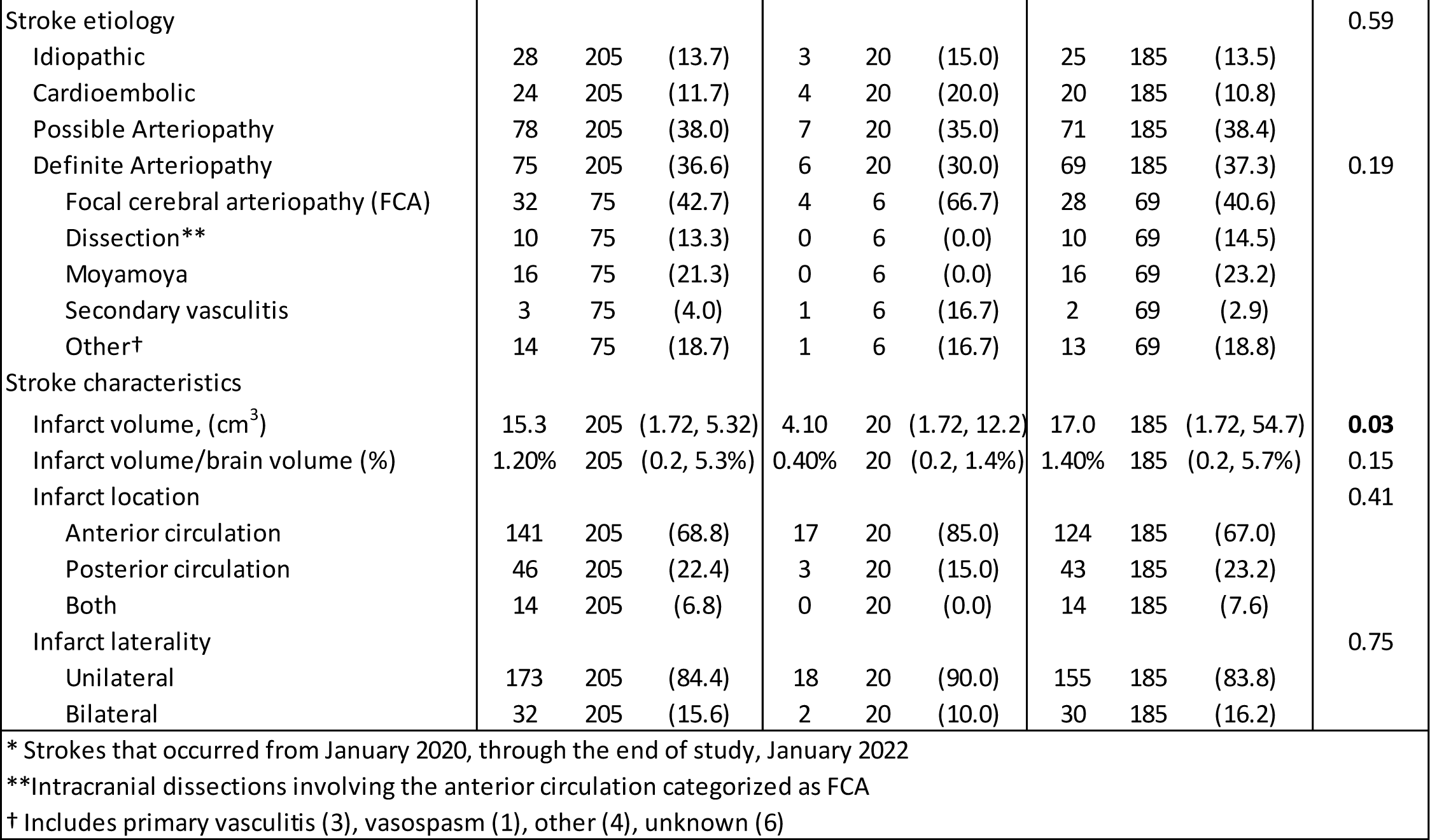
Characteristics of the 205 cases of childhood arterial ischemic stroke enrolled in the VIPS II study, stratified by serological evidence of recent VZV reactivation at the time of the stroke.

### Past VZV exposure (infection or vaccination)

Of the 205 cases, 198 (97%) had serological evidence of prior VZV exposure (IgG >0.1). Parents reported remote chicken pox in three (8.4, 10.6, and 11.7 years prior to the stroke), and prior VZV vaccination in 160 (78%). The median time from vaccination to stroke was 6.3 years (IQR 2.9-10.8; range 0.12-17.1). Age at the most recent VZV vaccination was a median of 4.3 years (IQR 1.6-5.3). All controls had serological evidence of prior VZV exposure. The parents reported remote chicken pox in five controls (5.3%) at a median (IQR) of 11.8 (8.3-14.4) years prior to enrollment, and prior VZV vaccination in 83 controls (87%) a median (IQR) of 7.2 (2.8-10.5) years prior to enrollment.

### Recent VZV reactivation

Positive baseline IgM titers were seen in 20/205 cases (9.8%; 95% CI 6.1-14.7%) and 3/95 controls (3.1%; 95% CI 0.66-8.9%) (p=0.06). All 20 cases and three controls had positive baseline IgG titers (>0.1) indicating past VZV exposure; hence, they had serological evidence of recent VZV reactivation (as opposed to primary infection). None had a recent vesicular rash to suggest cutaneous VZV infection (i.e., shingles). Elevated IgG titers provided additional support for an anamnestic response (**Figure 1**). Among the 20 positive cases, the median (range) IgG titer was 1.87 (0.24-3.31) absorbance units. Half of the cases (and none of the controls) had highly elevated IgG titers: IgG titers >2 in 10 cases (50%) and titers >3 in 6 cases (30%). The three positive controls had IgG titers <2: 0.13, 0.43, and 1.94.

**Figure 1.**
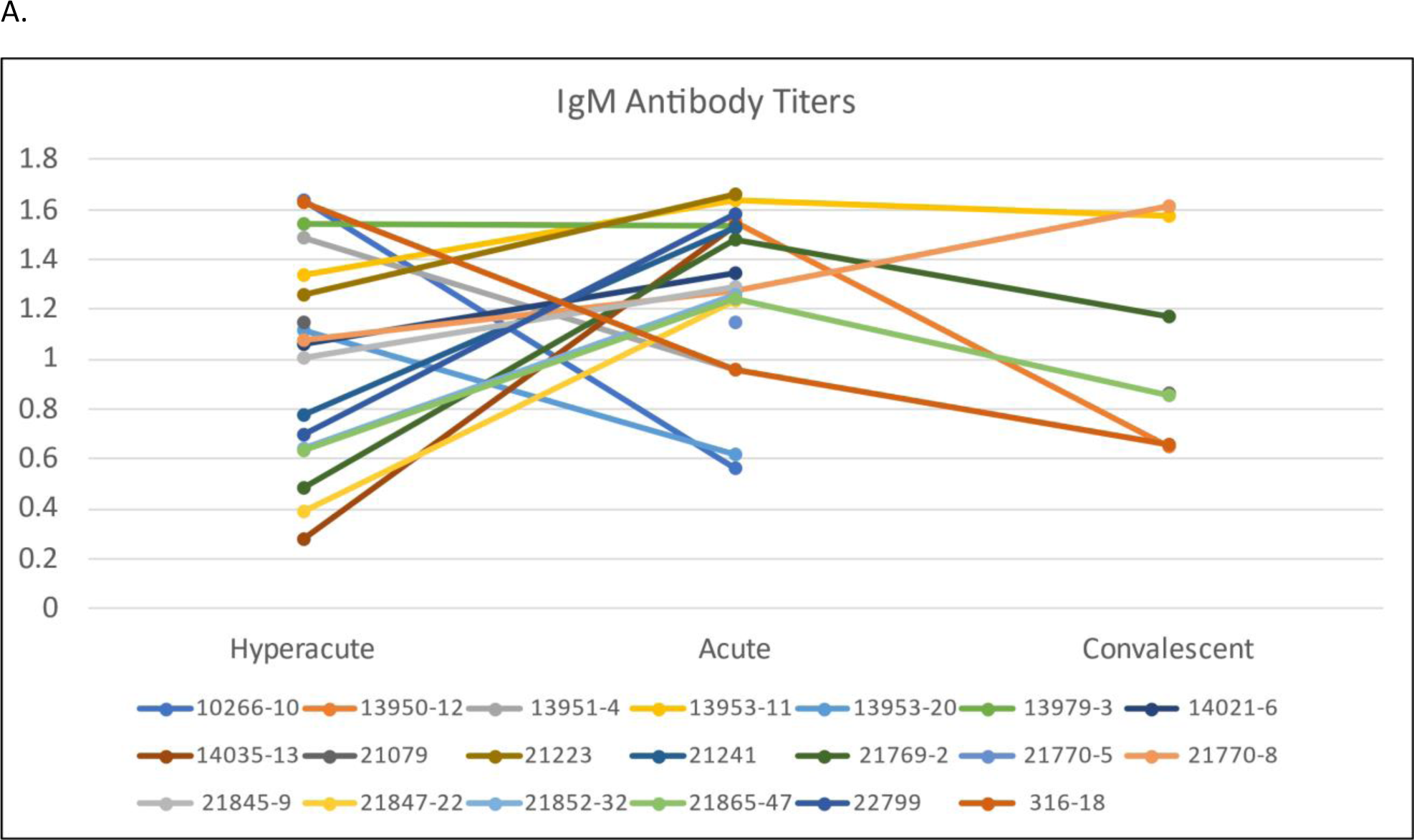
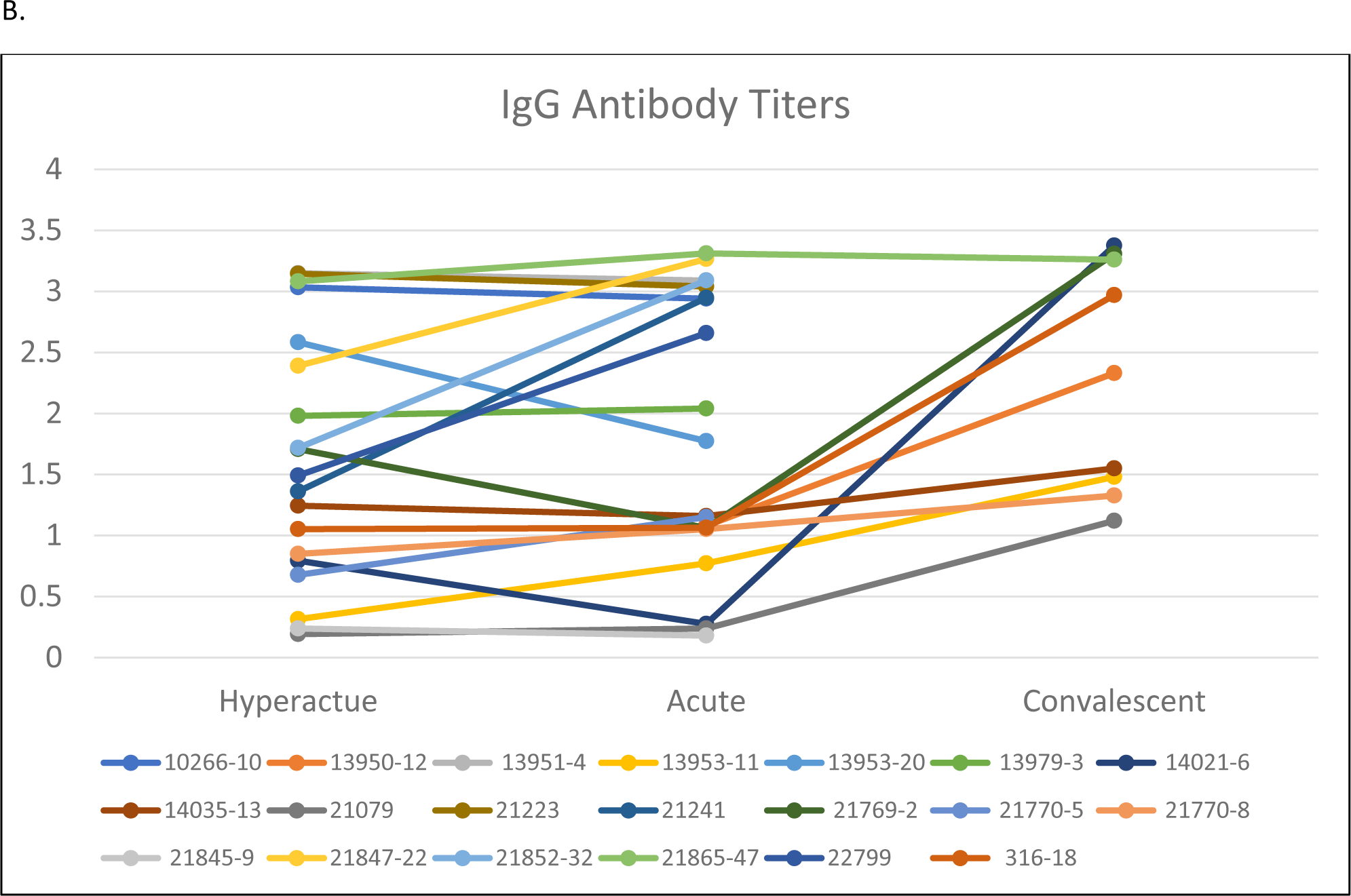
IgM (panel A) and IgG (panel B) VZV antibody titers at the hyperacute, acute, and convalescent time points in the 20 cases of stroke with serologic evidence of recent VZV reactivation at baseline. Y-axis is the antibody titer (absorbance units). Legend below the X-axis shows individual study IDs. A.

Parents of the 20 cases reported prior VZV vaccination in 16 and chicken pox in one (11.7 years prior); the remaining three had no reported VZV exposure. The median (range) time from vaccination to stroke was 6.1 (2.9-10.7) years for these 16 cases (**Figure 2**), similar to the median of 6.3 years for the full cohort.

**Figure 2.**
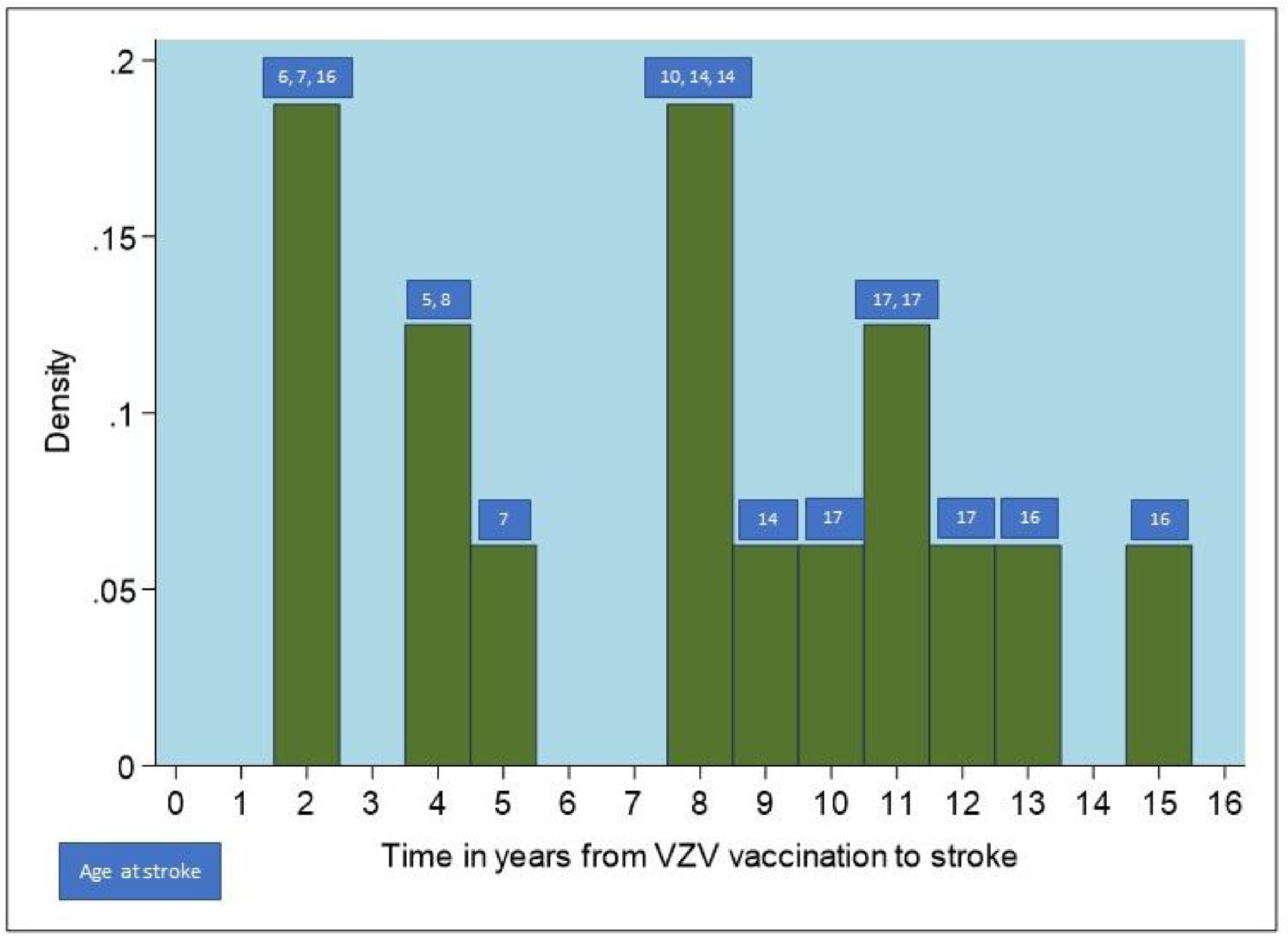
Distribution of time in years from the most recent VZV vaccination to stroke ictus among the 16 stroke cases with serologic evidence of recent VZV reactivation at baseline *and* prior VZV vaccination. Boxes above each bar show the age at stroke in years for the cases represented in that bar.

The 20 cases with recent VZV reactivation were similar to those without except for smaller infarct volumes (p=0.03) (**Table 1**). Of the 32 cases of definite FCA in the VIPS II cohort, 4 (12.5%) had recent VZV infection, compared to none of the definite cases of arterial dissection (n=10) or moyamoya (n=16), and only one of the 13 cases of definite arteriopathy that could not be further classified. However, evidence of recent VZV infection seen in all stroke subtypes: cardioembolic (4/24, 16.7%), idiopathic (3/28, 10.7%), and possible arteriopathy (7/78, 9.0%).

### VZV reactivation after stroke

Of the 74 cases with convalescent serum samples, 12 (16.2%, 95% CI 9-27%) had serologic evidence of VZV reactivation *after* the stroke: one had convalescent IgM titers ≥1.1, nine had rising IgG titers, and two had both. All were asymptomatic.

### VZV reactivation in well control patients

The aforementioned three well controls with elevated IgM titers were 11.0, 13.2, and 13.8 years of age. All had IgG titers indicating prior VZV exposure and had parental report of VZV vaccination (6 years prior, unknown timing, and 12 years prior, respectively). None had a history of chicken pox. In all three, the VZV reactivation was asymptomatic. When serum titers from all 95 controls were plotted, the distribution has a marked rightward skew suggesting that asymptomatic VZV reactivation can occur in well children (**Figure 3**).

**Figure 3.**
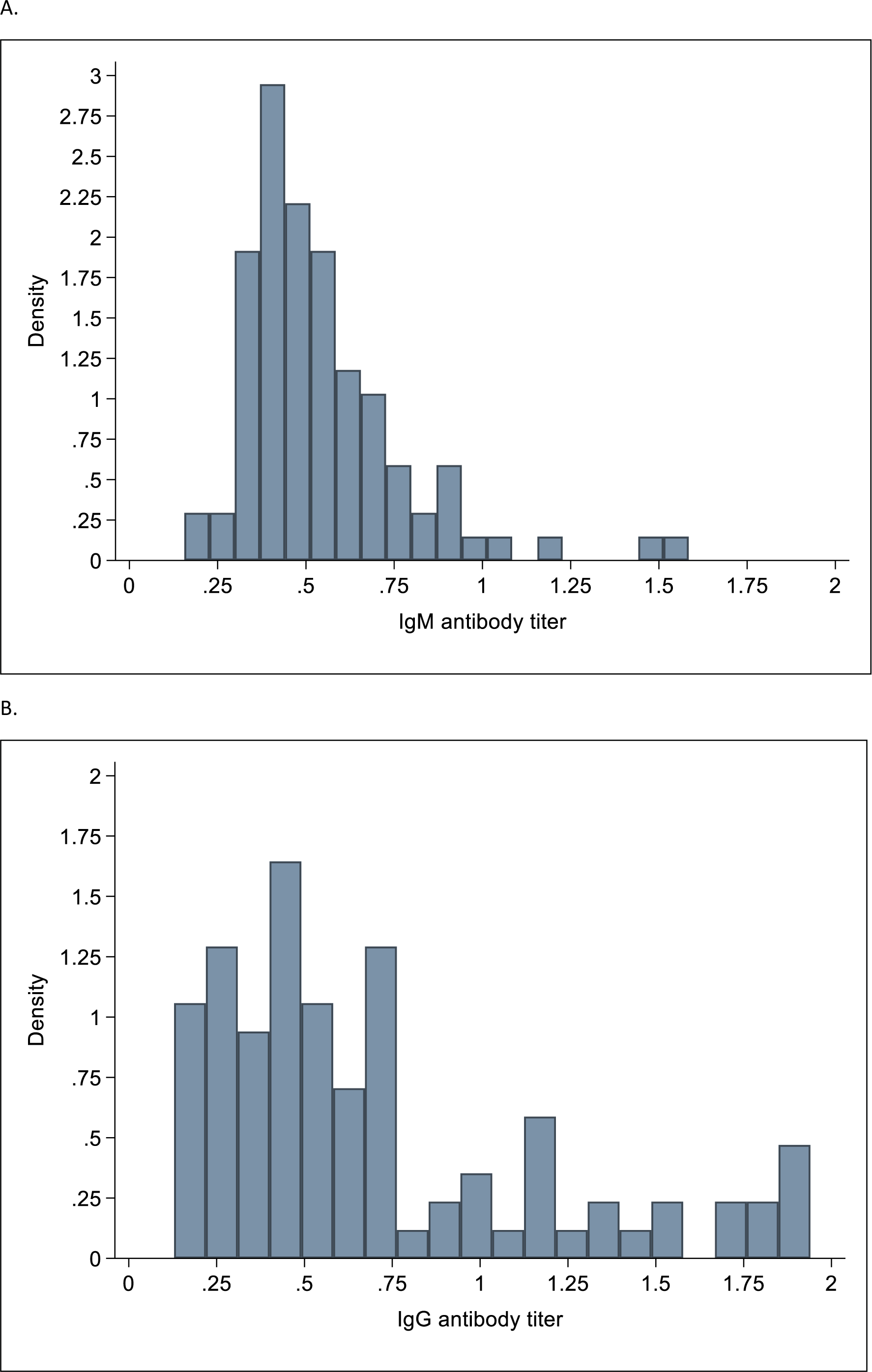
Distribution of IgM (panel A) and IgG (panel B) VZV antibody titers in 95 stroke-free well controls. Three controls had elevated IgM titers (>1.1) indicating recent VZV reactivation.

## Discussion

After primary VZV infection, which includes viremia and skin involvement (the characteristic vesicular rash of chicken pox), the adaptive immune system does not succeed in eradicating the virus.^20^ Rather, the virus enters sensory nerves and travels retrograde to the sensory ganglia (dorsal root and trigeminal nerve ganglia) where it becomes latent. Similarly, after childhood VZV vaccination, the live, attenuated vaccine virus first replicates in the skin, then travels to and enters a state of latency in the sensory ganglia.^21^ The latent period is dynamic: VZV antibody titers can rise with repeat exposure to exogenous virus, and the endogenous “latent” virus is thought to intermittently reactivate, triggering transient asymptomatic rises in VZV IgM antibodies.^20, 22, 23^ During periods of reduced immune function, VZV, whether wild-type or vaccine-derived, can reactivate and cause secondary infection, called zoster.^24^

In the VIPS II cohort of 205 children with spontaneous AIS, the vast majority had prior exposure to VZV, primarily through vaccination, and almost one in 10 had evidence of recent subclinical VZV reactivation. Their high IgM titers within days after the stroke suggest the recent reactivation *preceded* the stroke, supporting our original hypotheses that VZV reactivation may act as a stroke trigger even in the era of routine vaccination. An additional 16% of cases had serologic evidence of VZV reactivation *after* the stroke; while less important to our understanding of childhood stroke pathogenesis, this finding highlights the complex relationship between VZV and stroke and the need for careful interpretation of serologic data. The serologies in our well control children identified coincidental asymptomatic VZV reactivation in a small minority; this provides evidence supporting a dynamic latent phase with asymptomatic reactivation events occurring in the general vaccinated population of American children. (Some longitudinal studies of VZV vaccinated children may have missed these reactivation events because they did not measure IgM titers.^25^) The asymptomatic reactivation events in our childhood stroke cohort suggest that these events may not always be benign, but rather may be one of the more common viral stroke triggers in children. Contrary to our second original hypothesis, recent VZV reactivation was *not* more common in children with arteriopathic stroke compared to cardioembolic or idiopathic stroke. However, among definite arteriopathy cases, VZV reactivation was seen in FCA but not cervical dissection or moyamoya.

In the pre-vaccination era, multiple reports described a post-varicella arteriopathy (i.e., FCA) in previously healthy children with AIS following wild-type primary VZV infection (chickenpox).^8, 26, 27^ Rare autopsy studies revealed VZV particles and a lymphocytic infiltration in the arterial wall consistent with a focal angiitis.^28, 29^ These reports and an epidemiologic study from Canada indicated a delayed period of increased stroke risk occurring weeks to months after chicken pox.^30^ The virus travels within nerves at a rate of ≈10 cm/day, so the time interval between varicella and stroke is not solely explained by the time for virus to transit from the trigeminal ganglion to an artery.^31^ The time interval presumably includes the time required for virus within the artery to cause a sufficient inflammatory response to lead to stroke. However, to explain an interval of months, an additional latency period from primary infection to reactivation of the wildtype VZV in the trigeminal ganglion seems likely. In other words, many of these reported cases likely represented reactivation of wild-type VZV within months of the primary infection.

We enrolled the VIPS II cohort *after* VZV vaccination in young children became routine in North America and Australia. Japanese investigators first developed the live, attenuated vaccine in the 1970s by serially cultivating wild-type VZV in different cell cultures.^32^ The U.S. was the first country to approve the vaccine in 1995, with early policies recommending a single dose in children at 12-18 months and “catch-up” vaccination in susceptible children up to 13 years of age.^33^ In 2007, U.S. vaccine policy was revised to a two-dose schedule at 12-15 months and 4-6 years.^33^ Australia started a single-dose vaccination program in 2005, and Canadian provinces did the same between 2000 and 2007.^34, 35^ In the VIPS II cohort, the majority of parents reported prior vaccination (prior chicken pox in only three), and baseline IgG serologies supported prior VZV exposure in 97% of cases. The time interval from vaccination (initial VZV exposure) to stroke was generally prolonged—a median of six years—much longer than the historic time interval (weeks to months) between chicken pox and stroke. However, this prolonged time interval is similar to that reported in a small number of children with stroke after wild-type herpes zoster ophthalmicus.^36^ Eight children had chicken pox (initial VZV exposure) in early childhood and developed zoster (VZV reactivation) with stroke 2-12 years later.^36^

The asymptomatic VZV reactivation events in our cohort could represent reactivation of either vaccine-type virus or wild-type virus; our antibody tests were unable to distinguish the two. Case reports of vaccine-virus shingles in both immunocompetent and immunosuppressed children provide evidence that the vaccine virus remains latent in neurons (including the trigeminal ganglion) and can cause symptomatic reactivation.^24, 37^ A recent report describes 17 children and adolescents with zoster and meningitis caused by vaccine-type VZV; 14 of the 17 were immunocompetent.^38^ Half of the cases were adolescents, suggesting a multi-year delay after vaccination. It is plausible that subclinical vaccine-virus reactivation could have triggered stroke in some of the immunocompetent cases in our cohort.

Alternatively, the VZV reactivation events in our cohort could represent asymptomatic secondary infection with wild-type VZV in previously vaccinated children with waning immunity. In the VIPS I study, prior VZV vaccination significantly reduced risk of childhood AIS.^13^ Exposure to VZV vaccination is common North America and Australia, while exposure to wild-type VZV in children is now rare. However, children, including vaccinated children, can acquire a secondary wild-type varicella infection by exposure to an adult with herpes zoster (shingles), which occurs annually in over one million U.S. adults.^39, 40^ One adult with shingles can transmit the virus to up to eight other children and adults. The rarity of childhood AIS as an outcome in otherwise healthy children could be consistent with a rare secondary wild-type VZV infection after post-vaccination immunity has waned.

Based on the historical evidence of FCA after chicken pox (post-varicella arteriopathy), we hypothesized that VZV reactivation would more commonly precede arteriopathic stroke, particularly FCA, than cardioembolic or idiopathic stroke. Among the definite arteriopathy cases of our post-vaccination VIPS II cohort, recent VZV reactivation was indeed seen 12.5% of FCA cases and *not at all* in cases of the two other most common childhood arteriopathies (cervical dissection and moyamoya). Other neurotrophic pathogens have been linked to FCA,^41, 42^ including SARS-CoV-2;^43, 44^ and we anticipate that additional planned analyses of the VIPS II cohort will identify other infections in the FCA cases. However, we found recent VZV reactivation was not specific to arteriopathic stroke, with a similar prevalence in cardioembolic and idiopathic stroke. One potential explanation is misclassification of FCA cases. Early vascular imaging may be normal in FCA and many VIPS II cases lacked the serial imaging often needed to diagnose FCA; this could lead to misclassification as idiopathic. However, we observed a similar prevalence of recent VZV reactivation in the cardioembolic cases (16.7%), less likely to be misclassified cases of FCA.

More likely, VZV may act as a stroke trigger through multiple mechanisms that can contribute to all stroke subtypes. The mechanism hypothesized to cause FCA—reactivation of VZV latent in the trigeminal ganglion leading to an ICA arteriopathy —is presumably specific to that stroke subtype. However, the mechanisms by which viruses are generally postulated to trigger cerebrovascular and cardiovascular events are likely also relevant for VZV: circulating inflammatory cytokines affecting the coagulation system (“thromboinflammation”) and cardiac and arterial endothelium, favoring the formation of intracardiac or intraluminal thrombus.^45, 46^ Indeed, evidence in adults implicates VZV reactivation (shingles) as a trigger for both ischemic stroke and myocardial infarction, possibly through thromboinflammation or transient VZV-induced antiphospholipid antibodies.^47, 48^

The original VIPS I study enrolled patients from 2010 to 2014, and only 59% of childhood AIS cases reported prior VZV vaccination, compared to 78% of the VIPS II cohort (2016-2022).^5^ Fewer VIPS I cases had serologic evidence of past VZV infection (56% versus 97% in VIPS II), but the commercial antibody kits used in VIPS I, unlike the VIPS II assays, were insensitive for detecting low titers of IgG antibodies after VZV vaccination.^49^ However, we observed a similar prevalence of positive VZV IgM antibody titers at baseline in the two cohorts: 11.3% of VIPS I cases and 9.8% of VIPS II cases (**Table 2**).

**Table 2.** Comparison of VZV results in the VIPS I versus the VIPS II study.

| **Table 2.** Comparison of VZV results in the VIPS I versus the VIPS II study |  |  |  |  |  |  |
| --- | --- | --- | --- | --- | --- | --- |
| Study | Study Years | # of Cases | # VZV IgM positive | Percent VZV IgM positive | Clinical VZV | % Vaccinated |
| VIPS I | 2010-2014 | N=326 | n=37 | 11.3% | 0 | 58% |
| VIPS II | 2016-2022 | N=205 | n=20 | 9.8% | 0 | 78% |

Our study has a number of limitations. Because we have no pre-stroke serum samples, some misclassification of VZV reactivation as preceding versus following the stroke is possible. Our assumptions regarding the timing antibody titer elevations are based on other published studies with their own limitations.^17–19^ Our serologies could not differentiate vaccine-type versus wild-type VZV virus and we did not have CSF samples available for testing. However, prior studies have shown that many cases of wild-type VZV encephalitis do not have detectable VZV DNA in the CSF.^50^ The lack of serial vascular imaging, or standardized timing of imaging, is another major limitation and, as discussed above, likely led to under diagnosis of FCA. Almost 40% of all cases had “possible arteriopathy,” meaning that they lacked sufficient imaging and clinical data to confirm or exclude an arteriopathy. Of 75 cases of definite arteriopathy, 18% could not be further classified. The ongoing Focal Cerebral Arteriopathy Steroid (FOCAS) trial will address these limitations by enrolling possible cases of FCA, performing serial imaging at standardized time intervals, and measuring VZV serologies in the same research laboratory as VIPS II (ClinicalTrials.gov Identifier: NCT06040255).

Despite its limitations, the VIPS II study provides evidence that asymptomatic VZV reactivation events occur in childhood in association with AIS and, indeed, may be a particularly definable viral trigger for childhood AIS, present in about 10% of cases. Prior studies, including VIPS I, suggest that *many* childhood viruses may trigger stroke in children.^5, 13, 51, 52^ However, VZV is unique in several important ways: (1) many children are vaccinated against VZV; (2) VZV virus, whether wild-type or vaccine-type, remains latent in nerve ganglia, including the ganglion that innervates intracerebral arteries; (3) latent VZV can reactivate, and this reactivation may be clinical or asymptomatic, and benign or pathologic. Contrary to our expectations based on pre-vaccination pediatric stroke literature, the role of VZV in the VIPS II cohort was not limited to cases of FCA; however, adult vascular literature supports VZV reactivation as a trigger for stoke and myocardial infarction, suggesting a broader pathologic role for VZV. There remains a conundrum regarding the role of vaccine virus versus wild-type virus. Although vaccinated children may silently contract wild-type VZV from adults with herpes zoster, the prevalence of this remains unknown;^40^ hence, we cannot speculate which type of VZV is more likely reactivating in our cohort. Because VZV vaccination protects against childhood stroke, among its other benefits,^13, 40^ waning immunity could explain the persistent ability for VZV to trigger childhood stroke in vaccinated children.

## Data Availability

The date are available from the authors upon request.

## ACKNOWLEDGMENTS

The authors wish to acknowledge the important contributions of the research coordinators at VIPS II sites, the VIPS II project managers, Maria Kuchherzki and Kathleen Colao, the virology laboratory manager, Wallen Jackson, and the patients and their families. We also acknowledge Maria Kuchherzki for helping to create the graphic abstract.

## VIPS II Investigators (in order of enrollment)

Michael Dowling, Christine Fox, Nomazulu Dlamini, Gabrielle DeVeber, Marcela Torres, Jenny Wilson, Sarah Lee, Dana Cummings, Warren Lo, Melissa Chung, Lori Jordan, Tim Bernard, Megan Barry, Rebecca Ichord, Lauren Beslow, Mukta Sharma, Shannon Carpenter, Catherine Amlie-Lefond, Neil Friedman, John Michael Taylor, Michael Rivkin, Laura Lehman, Paola Pergami, Andrea Pardo, Tracee Ridley-Pryor, Ryan Felling, Lisa Sun, Mark Mackay, Adam Kirton.

---

**Supplemental Table 1.** Characteristics of the 205 cases of childhood arterial ischemic stroke and 95 stroke-free well controls with blood samples in the VIPS II study.

| Characteristic | Cases<br>(N=205) |  |  | Well Controls<br>(N=95) |  |  | p-value |
| --- | --- | --- | --- | --- | --- | --- | --- |
|  | n | N | (%) | n | N | (%) |  |
| **Demographics** |  |  |  |  |  |  |  |
| Age in years, median (IQR) | 11.6 | 205 | (5.5, 15.6) | 11.8 | 95 | (6.8, 15.3) | 0.59 |
| Sex |  |  |  |  |  |  | 0.69 |
| Female | 92 | 205 | (44.9) | 45 | 95 | (47.4) |  |
| Male | 113 | 205 | (55.1) | 50 | 95 | (52.6) |  |
| Race/ethnicity |  |  |  |  |  |  | 0.005 |
| White, non-Hispanic | 111 | 205 | (54.1) | 48 | 95 | (50.5) |  |
| Black, non-Hispanic | 25 | 205 | (12.2) | 9 | 95 | (9.5) |  |
| Asian | 15 | 205 | (7.3) | 5 | 95 | (5.3) |  |
| Hispanic | 44 | 205 | (21.5) | 15 | 95 | (15.8) |  |
| Native American/First Nations | 1 | 205 | (0.5) | 0 | 95 | (0.0) |  |
| Mixed or other | 8 | 205 | (3.9) | 13 | 95 | (13.7) |  |
| Unknown | 1 | 205 | (0.5) | 5 | 95 | (5.3) |  |
| **Socioeconomic Status** |  |  |  |  |  |  |  |
| Residence |  |  |  |  |  |  | 0.03 |
| Urban | 39 | 204 | (19.1) | 24 | 95 | (25.3) |  |
| Suburban | 116 | 204 | (56.9) | 60 | 95 | (63.2) |  |
| Rural | 49 | 204 | (24.0) | 11 | 95 | (11.6) |  |
| Household income (in US dollars) |  |  |  |  |  |  | 0.17 |
| <\$10,000 | 13 | 205 | (6.3) | 4 | 94 | (4.3) | |
| \$10,000-30,000 | 24 | 205 | (11.7) | 8 | 94 | (8.5) | |
| \$31,000-50,000 | 26 | 205 | (12.7) | 11 | 94 | (11.7) | |
| \$50,000-100,000 | 45 | 205 | (22.0) | 24 | 94 | (25.5) | |
| >100,000 | 59 | 205 | (28.8) | 47 | 94 | (50.0) |  |
| Maternal education, highest level |  |  |  |  |  |  | 0.001 |
| Less than high school | 15 | 202 | (7.4) | 1 | 95 | (1.1) |  |
| High school graduate | 46 | 202 | (22.8) | 14 | 95 | (14.7) |  |
| Some college education | 60 | 202 | (29.7) | 20 | 95 | (21.1) |  |
| Bachelor's degree, or equivalent | 40 | 202 | (19.8) | 26 | 95 | (27.4) |  |
| Some graduate education | 8 | 202 | (4.0) | 1 | 95 | (1.1) |  |
| Graduate degree | 33 | 202 | (16.3) | 33 | 95 | (34.7) |  |

